# Randomized, Crossover Clinical Trial on the Safety, Feasibility, and Usability of the ABLE Exoskeleton: A Comparative Study with Knee-Ankle-Foot Orthoses

**DOI:** 10.1101/2023.04.11.23288209

**Authors:** Antonio Rodríguez-Fernández, Joan Lobo-Prat, Mariona Tolrà-Campanyà, Florentina Pérez-Cañabate, Josep M. Font-Llagunes, Lluis Guirao-Cano

## Abstract

Wearable exoskeletons are emerging as a new tool for gait training. However, comparisons between exoskeletons and conventional orthoses in terms of safety and feasibility are scarce. This study assessed the safety, feasibility, usability, and learning process of using the ABLE Exoskeleton in people with spinal cord injury (SCI) while comparing it with knee-ankle-foot orthoses (KAFOs). In this randomized, crossover clinical trial, 10 patients with chronic complete SCI (T4-T12) conducted a 10-session training and assessment protocol with each device: KAFOs and the ABLE Exoskeleton. Outcomes on safety (adverse events), and feasibility and usability (level of assistance, donning/doffing, therapy activities) were recorded for both devices. Evaluation sessions included standard clinical tests (Timed Up and Go, 10-Meter Walk Test, and 6-Minute Walk Test) to assess gait performance. The therapy metrics (number of steps, distance, gait speed, and standing and walking time) were recorded at each session for the robotic device. Participants quickly learned how to use the ABLE Exoskeleton, showing improvements in all therapy metrics (p*<*0.05) and the 6-Minute Walk Test (p*<*0.05). The robotic device reported less adverse events than KAFOs (17 and 31, respectively). Total donning and doffing time was 43 seconds faster with the robotic device using comparable levels of assistance. The time to complete the therapy activities was very similar between devices. Overall, participants needed 1 to 4 training sessions to perform essential therapy activities (sit/stand transitions, walking 10 meters, turning around) with both devices using minimum assistance or less. The results of this study show that it is feasible and safe for people with motor complete paraplegia due to SCI (T4-T12) to use the ABLE Exoskeleton for gait training in a rehabilitation hospital setting. The ABLE Exoskeleton proved to be safer than KAFOs in terms of adverse events, and as practical and easy to use as the conventional orthoses.

## Introduction

Spinal cord injury (SCI) is a life-changing condition that results in sensory and motor impairments that are often associated with permanent paralysis of the lower limbs [1]. Therefore, recovering the ability to stand and walk independently has a significant impact on participation in social and professional activities [2, 3], along with general health and well-being [4–6]. For many years passive orthoses, such as knee-ankle-foot orthoses (KAFOs) –which are the current standard of care for verticalization and gait ambulation in people with SCI–, have served that purpose. However, using these orthoses can often be challenging and inconvenient for the user [7–10].

Gait training using rehabilitation robotic technology has expanded rapidly in the last years due to its advantages over conventional therapy: it increases the duration and intensity of sessions while performing more accurate and continuous physiological movements, and reduces the physical loads of the therapists [11–14]. Since their first appearance in the clinical setting 25 years ago [15], several rehabilitation robots for gait training have been developed which can broadly be classified into grounded exoskeletons, grounded end-effectors, and wearable exoskeletons [16]. In the last years, robotic gait rehabilitation for people with SCI is evolving principally towards wearable exoskeletons, since they promote more active participation of the user than grounded robots and allow ambulation in the community setting [6].

The safety and feasibility of wearable exoskeletons for gait rehabilitation after SCI have been evaluated in previous studies [3, 17–24]. In fact, a number of exoskeletons have already been certified for use in the clinical setting [6] or even at home [25, 26]. However, comparisons supporting their superiority over conventional passive orthoses to assist locomotion are still scarce. Only a few studies aimed to compare wearable exoskeletons and passive orthoses; however, their main focus was on the comparison of functional performance, energy consumption, and/or patient satisfaction using each of the two systems [27–33]. Thus, the safety, feasibility, and usability of wearable exoskeletons have never been compared to those of passive orthoses, such as KAFOs. Consequently, it is unknown how far wearable exoskeletons differ from conventional passive orthoses in these regards.

In like manner, despite the technological development and the satisfactory acceptance that wearable gait-assistive exoskeletons are having, intensive and long training is necessary to learn to use these devices independently [34–36]. In addition, the number of sessions and the skill needed to control a wearable exoskeleton differs among users and it depends on different aspects such as level of injury (LOI), body mass index (BMI), age, and lifestyle [37]. Various studies tested the use of wearable exoskeletons in people with SCI to perform different tasks [34–40]. However, all the preceding studies assessed hip-knee-powered exoskeletons, and only one used a knee-powered exoskeleton [24]. The latter examined the use of the ABLE Exoskeleton in people with SCI, both with motor complete and incomplete injuries, and primarily in the acute or subacute phase (i.e., onset of paralysis injury within the last year). As a result, little is known about how difficult it is for people with SCI in the chronic phase to operate and learn to use a knee-powered exoskeleton.

We conducted a randomized, crossover clinical trial comparing the use of conventional KAFOs against a robotic knee-powered lower limb exoskeleton (i.e., the ABLE Exoskeleton) for gait training in people with SCI. The primary outcome measure of the clinical trial was the metabolic cost of walking using both devices, which was already assessed in [33]. The main objective of the present study was to evaluate the safety, feasibility, and usability of the ABLE Exoskeleton in a rehabilitation clinical setting. The secondary objectives were to compare the ABLE Exoskeleton with conventional KAFOs in terms of safety, feasibility, and usability; and to gain insight into the learning process of using the ABLE Exoskeleton.

## Materials and methods

### Study design

A randomized, crossover clinical trial was conducted to compare gait training with a knee-powered bilateral lower limb exoskeleton (i.e., the ABLE Exoskeleton) against KAFOs (i.e., the standard of care for verticalization and ambulation in people with SCI). The clinical trial was performed at Asepeyo Sant Cugat Hospital (Barcelona, Spain), a center specialized in SCI, from February to August 2021. The time to complete the study for each patient covered approximately 12 weeks. All the participants were randomized to one of the groups: KAFO or ABLE. The principal investigator blindly chose one of the 20,000 lines of a book of random numbers [41]. The 10 first numbers of the chosen line (from left to right) were used to allocate each of the participants to one of the two groups following the order of enrollment: even numbers assigned participants to the ABLE group and odd numbers assigned participants to the KAFO group.

The clinical trial (study code: 2020/157-REH-ASEPEYO) was approved by the responsible ethics committee (CEIm Grupo Hospitalario Quirónsalud-Catalunya) and the national competent authority (Spanish Agency of Medicines and Medical Devices (AEMPS), EUDAMED: CIV-ES-21-01-035724). The study conformed to the principles of the Declaration of Helsinki (revised version 2013), the ISO 14155:2011, and the European Regulation MDR 2017/745 on medical devices. The study protocol was first registered at ClinicalTrials.gov on 25/03/2021 (NCT04855916). Note that due to organizational issues, the investigation team failed to register the study before the first participant enrollment. As a result, five participants had been enrolled before registration. Nonetheless, S1 File shows the Clinical Research Ethics Committee’s resolution of the trial protocol, which was accepted prior to the study’s start date and has not been modified since then; demonstrating the clinical trial’s prospective nature.

### Participants

Eleven outpatients from the investigational site with chronic (i.e., time since injury more than one year ago) motor-complete SCI (AIS grade A/B) were assessed for eligibility in this study. One patient was excluded for not meeting the inclusion/exclusion criteria (see S2 Table) and 10 were enrolled (Fig 1) and completed the entire protocol (Table 1). The neurological LOI of the enrolled participants ranged from T4 to T12. Participants were 44.10 ± 5.93 years old and mostly male (9 out of 10). Seven participants had a traumatic SCI, while three had a non-traumatic SCI. All participants had previous experience using KAFOs, and three of them had previously used other lower extremity wearable exoskeletons. Prior to data collection, patients provided written informed consent to take part in the study. During the clinical trial, the medical team, whose members are co-authors of the present publication, had access to the participants’ information.

**Table 1.** Patient demographics.

|  | Gender | Age range | Level of injury | Time since injury | AIS [42] | Height (cm) | Weight (kg) | Starting device |
| --- | --- | --- | --- | --- | --- | --- | --- | --- |
| **P1** | M | 36-40 | T4 | 11 | A | 176 | 80 | K |
| **P2** | F | 46-50 | T4 | 7 | A | 168 | 70 | K |
| **P3** | M | 41-45 | T4 | 12 | A | 170 | 74 | A |
| **P4** | M | 36-40 | T6 | 21 | A | 174 | 72 | A |
| **P5** | M | 51-55 | T8 | 5 | A | 170 | 80 | A |
| **P6** | M | 46-50 | T11 | 2 | A | 183 | 77 | K |
| **P7** | M | 31-35 | T12 | 8 | A | 169 | 84 | K |
| **P8** | M | 46-50 | T8 | 10 | B | 173 | 75 | A |
| **P9** | M | 46-50 | T11 | 23 | A | 185 | 98 | A |
| **P10** | M | 41-45 | T10 | 6 | B | 173 | 71 | K |

AIS: ASIA (American Spinal Injury Association) Impairment Scale; M: Male, F: Female, K: KAFOs, A: ABLE Exoskeleton.

**Fig 1.**
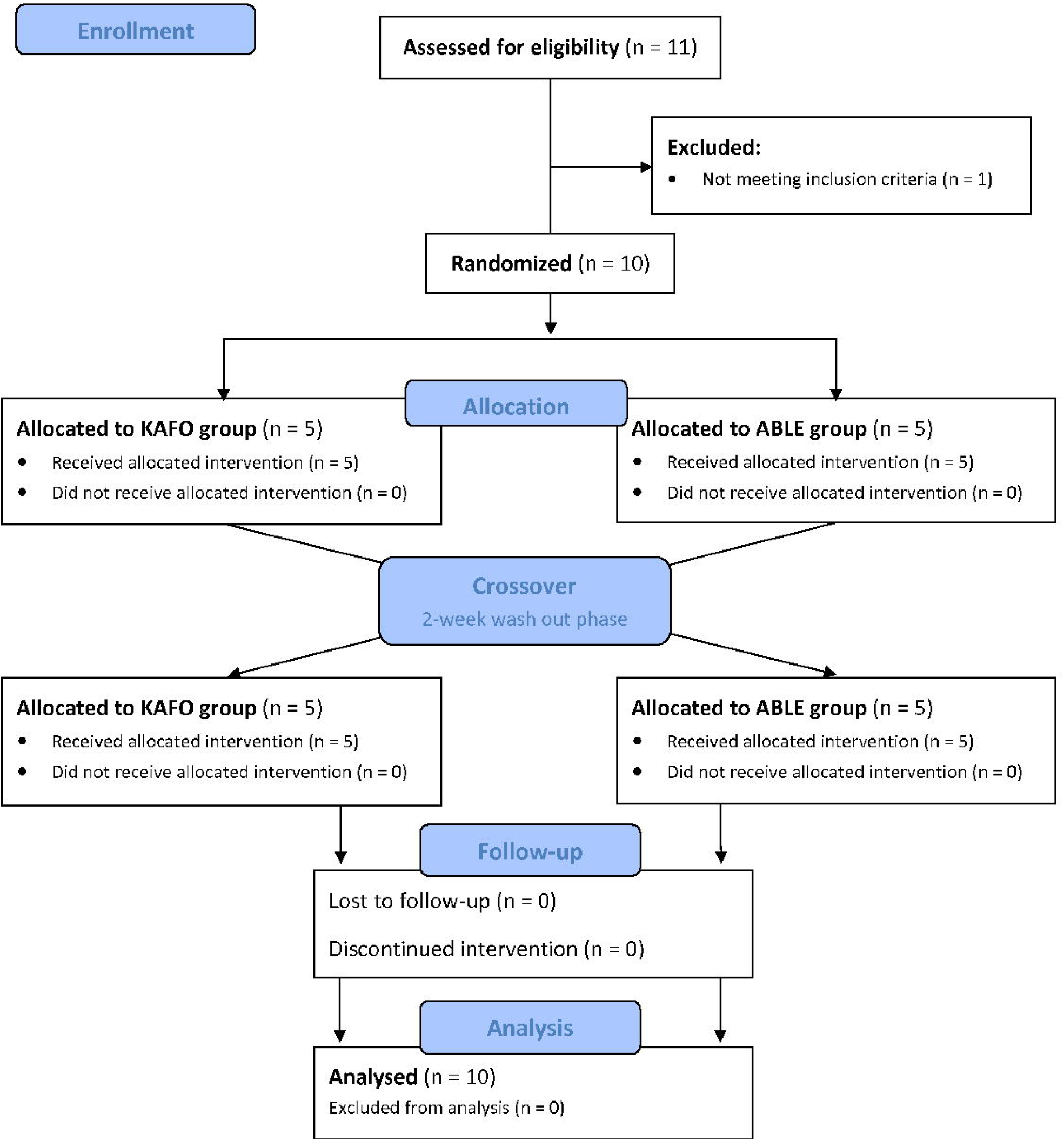
CONSORT flow diagram

### Study protocol

Participants in both groups attended training sessions of 90 minutes of duration twice a week for five consecutive weeks, which resulted in a total of 10 sessions: eight overground gait training sessions (sessions 1 to 4 and 6 to 9) and two evaluation sessions (sessions 5 and 10). Once all the sessions were completed, and after a two-week resting period, participants repeated the same process with the crossed-over device. The training sessions were carried out by at least one trained therapist plus an additional therapist or assistant if required. During each of the overground gait training sessions, participants spent a minimum of 30 minutes doing sit-to-stand and stand-to-sit transitions, and standing and walking exercises using one of the two devices and the aid of a walker. The training sessions were scheduled for 90 minutes to also include adjustments, donning and doffing time, and data collection time. Moreover, participants performed different therapy activities (grouped by balance abilities, walking abilities, and advanced abilities) that increased in difficulty (Table 2) and were adapted from [36].

**Table 2.** Therapy activities.

| Ability | Number | Activity description |
| --- | --- | --- |
| Balance | 1 | Sit-to-stand |
|  | 2 | Weight shifting |
|  | 3 | Manipulate remote controller while standing (only for ABLE Exoskeleton) |
|  | 4 | Touch the head while standing |
|  | 5 | Stand-to-sit |
| Walking | 6 | Walk 10m (with stops) |
|  | 7 | Start and stop walking with dominant leg |
|  | 8 | Start and stop walking with non-dominant leg |
|  | 9 | Walk 10m (without stops) |
|  | 10 | Turn around |
| Advanced | 11 | Walk close to a chair, turn around and sit down |
|  | 12 | Walk a 90° turn to the right (with stops) |
|  | 13 | Walk a 90° turn to the right (without stops) |
|  | 14 | Walk a 90° turn to the left (with stops) |
|  | 15 | Walk a 90° turn to the left (without stops) |
|  | 16 | Walk close to an object, stop and manipulate it |
|  | 17 | Walk in an arrow area |
|  | 18 | Stop in front of a door, open it (outward), and continue walking |
|  | 19 | Stop in front of a door, open it (inward), and continue walking |
|  | 20 | Stop near a wall, turn, and lean against it |
|  | 21 | Walk on different surfaces (e.g., yoga mat, uneven terrain) |
|  | 22 | Walk a slalom |

In the evaluation sessions, participants performed three standardized clinical tests using each of the two devices and the help of a walker: the Timed Up and Go (TUG), the 6-Minute Walk Test (6MWT), and the 10-Meter Walk Test (10MWT). Participants started with the TUG, which consisted of walking back and forth in a three-meter pathway, starting and finishing seated on a chair. Before and after completing the TUG, participants remained in a sitting position for three minutes. This test was repeated twice with a five-minute break in-between to recover from fatigue. The 6MWT was conducted after a resting period of 15-20 minutes after the second TUG. Before the 6MWT, participants were asked to rest in a sitting position for three minutes and, 30 seconds before the end of this resting period, they were asked to stand up and get ready to start. During the test, participants walked in a 10-meter pathway at a comfortable speed selected by themselves. Participants were allowed to rest and stop the test as necessary. The 10MWT was measured during the first 10 meters of the 6MWT.

After the final evaluation session (i.e., session 10), participants answered the Quebec User Evaluation of Satisfaction with Assistive Technology (QUEST 2.0) [43] and the Psychosocial Impact of Assistive Devices Scale (PIADS) [44] questionnaires to evaluate users’ satisfaction and psychosocial impact, respectively, that the training with the corresponding device may have had. Results from the questionnaires were presented previously in [33]. Finally, four weeks after the final evaluation session, a follow-up phone call was conducted to monitor any issues that may have occurred.

### Gait assistive devices

#### ABLE Exoskeleton

The ABLE Exoskeleton (ABLE Human Motion S.L., Barcelona, Spain) is a knee-powered lower limb exoskeleton that actively assists a person to stand up, walk and sit down (Fig 2a). It consists of a rigid brace that attaches to the user’s lumbar region, thighs, shanks, and feet via textile straps and rigid supports. The exoskeleton assists with knee flexion-extension movements through battery-powered actuators and allows free hip flexion-extension movements via a passive hinge joint. The exoskeleton size can be adjusted on the length and width of the shank and thigh segments and the hip-width to fit users with a height of 150-190 cm and a maximum weight of 100 kg. The battery is placed on the lumbar module and the total mass of the device is 9.80 kg.

**Fig 2.**
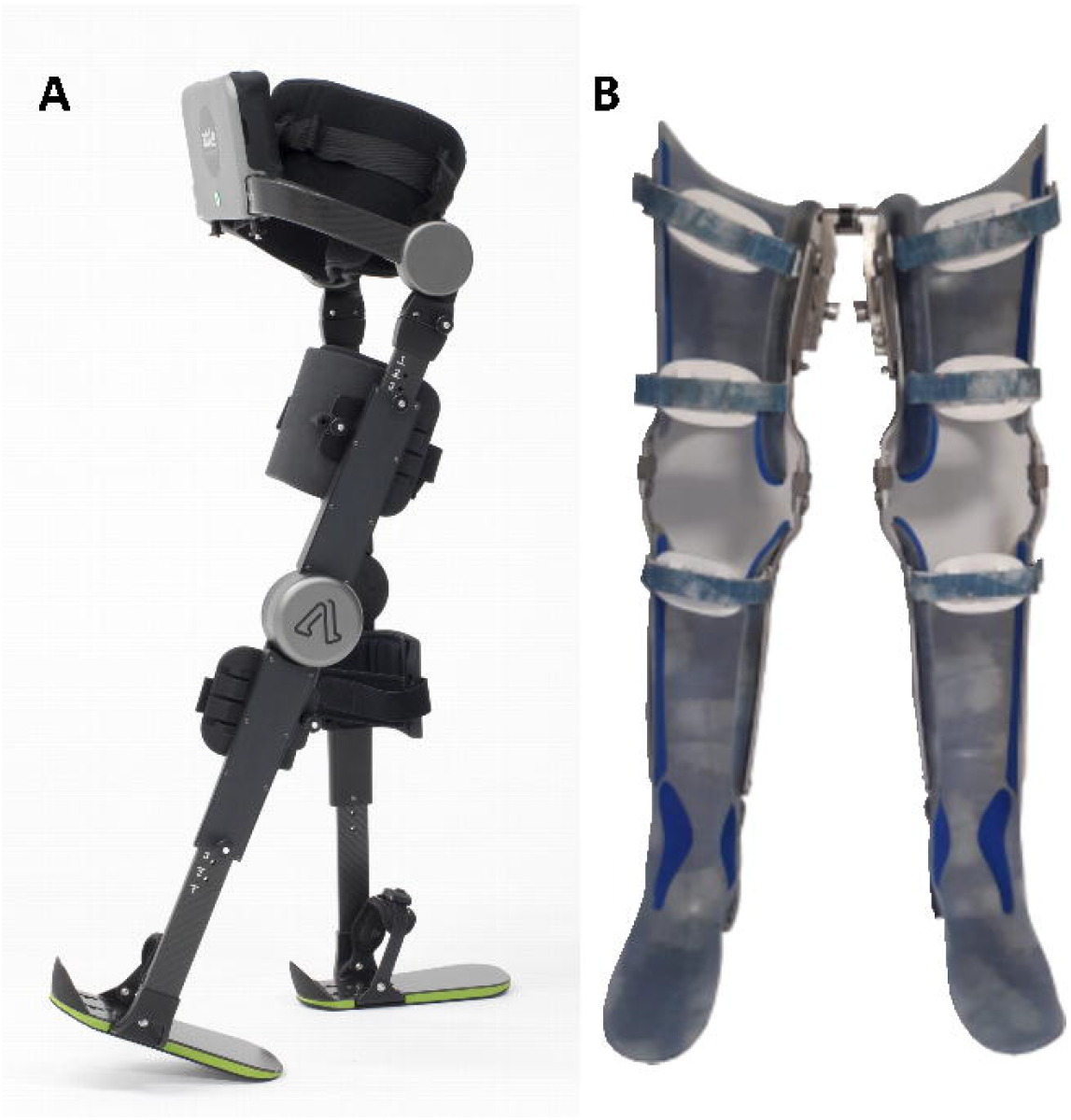
(**a**) Knee-powered lower limb exoskeleton; i.e., the ABLE Exoskeleton (ABLE Human Motion S.L., Barcelona, Spain). (**b**) Knee-ankle-foot orthosis with Walkabout.

The initiation of each step can be activated manually by the therapist (push-buttons at the lumbar module) or automatically by the patient detecting their intention to step forward, the latter by measuring a change in the thigh angular velocity with an inertial measurement unit (IMU). The device comes with a smartphone with the pre-installed software application ABLE Care (ABLE Human Motion S.L., Barcelona, Spain) that allows the therapist to configure the step parameters and monitor the exoskeleton. In addition, the ABLE Exoskeleton presents two operation modes to transition between states (i.e., sitting-to-standing, standing-to-walking, walking-to-standing, and standing-to-sitting) based on the user’s expertise. For beginners, the exoskeleton can be controlled by the therapist using the lumbar buttons or ABLE Care. For more advanced users, state transitions can be controlled by the users themselves through a remote controller attached to the walker.

#### Knee-ankle-foot orthoses

Knee-ankle-foot orthoses are passive leg braces personalized to each individual that provide stability by locking the knee joint at full extension and the ankle at the neutral position [8]. In general, it consists of an orthopedic thermoplastic cast of the thigh and shank segments attached to metal bars that are connected to a foot insole (Fig 2b).

All the participants in this study used their own KAFOs made of thermoplastic that had been prescribed from the investigational site. Note that eight out of the 10 participants used the KAFOs in combination with a multiaxial subperineal hip joint (also known as Walkabout joint [45, 46]), that stabilizes stance and provides a reciprocating gait. The average mass of the KAFOs including the Walkabout unit was 3.16 kg (ranging from 2.5 to 3.7 kg).

### Data collection

The primary endpoints for evaluating the safety of the devices (i.e., the ABLE Exoskeleton and KAFOs) were the number of adverse events (AEs), serious adverse events (SAEs), and study drop-outs related to the use of the device. SAEs and AEs were classified systematically according to ISO 14155:2011, MEDDEV 2.7/3, and MDCG 2020-10/1. AEs were defined in four categories: (1) device-related (AEs that have occurred as a direct result of the device itself), (2) procedure-related (AEs that occurred as a result of the activities performed in training, but were not caused by the device), (3) disease-related (AEs that occurred as a result of the underlying health conditions), and (4) other causes or undetermined relation. Finally, device deficiencies without a medical occurrence were also registered.

Additionally, specific AEs monitoring included: (1) checks for skin lesions pre- and post-session following the European Pressure Ulcers Advisory Panel (EPUAP) scale [47], (2) assessment pre- and post-session of presence, location, and severity of pain using a 0-10 visual analogue scale (VAS), and (3) documentation of falls or any event that required medical intervention.

The primary endpoints for feasibility and usability were the level of assistance (LOA) and gait performance during the standardized clinical tests (i.e., time to complete the TUG, distance covered during the 6MWT, and gait speed and cadence in the 10MWT), the number of sessions required to accomplish the therapy activities, and the time and LOA needed to don and doff the devices. The donning period began when the participant was ready to transfer from the wheelchair to the stretcher/chair where the device was placed and ended when the device was completely adjusted. The doffing period started when the participant was ready to take off the device until transferred back to the wheelchair. In addition, and only for the ABLE Exoskeleton, the participants’ ability to use the robotic device was assessed by the therapy metrics recorded by the device: number of steps, distance, gait speed, therapy time, and standing and walking time. Note that therapy metrics started to be recorded when the therapist initialized the training session through the mobile app ABLE Care (i.e., generally after donning the exoskeleton) and finished when the therapist ended up the session (i.e., generally before doffing the exoskeleton).

The LOA was recorded in each session for donning/doffing the devices, and in sessions 5 and 10 for each of the clinical tests. The LOA was reported by the clinical staff using a rating scale adapted from the Functional Independence Measure (FIM) [48]. The only difference with the FIM was that the modified independence score (i.e., the patient requires the use of a device, but no physical assistance) was removed since all the participants in this study used an assistive device for walking (Table 3).

**Table 3.** Level of Assistance (LoA) definitions for the process of donning/doffing the device and for the therapy activity tasks and standardized clinical tests.

| Level of assistance | Donning/doffing | Therapy activity task |
| --- | --- | --- |
| Total assistance | Participant performs 0-25% of the effort to don/doff the device. Participant is essentially reliant on the trainer to perform all aspects of the donning/doffing | Participant performs 0-25% of the effort to use the device. Two therapists are required to support the participant in the device at all times |
| Maximum assistance | Participant performs 25-50% of the effort to don/doff the device. Participant needs maximum assistance to transfer to device and position legs, but may be able to adjust the thigh straps | Participant performs 25-50% of the effort to use the device. Participant needs maximum assistance from the therapist to remain balanced |
| Moderate assistance | Participant performs 50-75% of the effort to don/doff the device. Participant needs moderate assistance to transfer to device and position legs, but may be able to adjust the thigh and shin straps | Participant performs 50-75% of the effort to use the device. The therapist has both hands on the participant or device at all times to provide occasional guidance or balance support |
| Minimal assistance | Participant performs >75% of the effort to don/doff the device. Participant can transfer to device and adjust straps, but may need help to position legs | Participant performs >75% of the effort to use the device. The therapist has one hand on the participant or device for infrequent guidance or balance |
| Supervision | The therapist is not touching the participant, but may provide verbal prompts or contact guarding to ensure safety | The therapist is not touching the participant, but is close enough to provide support for balance or guidance as needed |
| Independence | Participant is fully independent donning/doffing device | Participant is fully independent while using the device and the therapist does not provide any assistance |

The time to complete the TUG was measured from the start of the test until the participant sat down on the chair. For the 6MWT, the distance covered was recorded from the start of the test until the time of the test finished or the participant ceased the test and sat down on the chair. The gait speed and cadence during the 10MWT were calculated from the middle-placed six meters of the 10-meter pathway (the first and last two meters were taken for acceleration and deceleration, respectively).

The number of sessions needed to accomplish each of the therapy activities for the first time was recorded by the therapists. The activity was considered as successfully completed if the participant performed the activity with a LOA of *Minimum assistance, Supervision*, or *Independence*. The list of the therapy activities was ordered in increasing complexity as shown in Table 2.

Finally, to analyze the learning process of using the ABLE Exoskeleton, we studied the time to complete the ability activities (Table 2), adapted from [36], and the evolution of the therapy metrics recorded by the device.

### Data and statistical analysis

The sample size of the present clinical study (i.e., 10 patients) was selected taking into account previous clinical investigations that also had the purpose of analyzing the performance of a robotic exoskeleton against passive orthoses (e.g., KAFOs) [27–29, 31].

The AEs and SAEs were described using group sizes and frequencies. Quantitative variables were summarized using standard descriptive statistics (i.e., mean, standard deviation (SD), minimum, and maximum).

Statistical analysis of feasibility and usability metrics considered all the study participants. Note that P10 missed the first training session with the ABLE Exoskeleton and, therefore, we considered the second training session of P10 as it was the first one for baseline comparisons.

For metrics assessing the learning process of using the ABLE Exoskeleton, we analyzed the data to look for outliers, which were confirmed using the MATLAB *isoutlier* function (i.e., an outlier is a value that is more than three scaled median absolute deviations away from the median). Moreover, evaluation sessions (i.e., sessions 5 and 10) were not included in the statistical analysis due to the differences in the protocol with respect to the gait training sessions. Finally, the LOA and the therapy activities metrics were qualitatively compared.

The differences between devices and sessions were analyzed using paired two-tailed t-tests or Wilcoxon signed-rank tests based on the distribution’s normality quantified by the Kolmogorov–Smirnov statistic. The level of significance was set to *α* = 0.05. Statistical analyses were carried out using R version 4.2.0.

## Results

The 10 study participants completed the protocol without deviations, with only P10 missing one training session (session 1 with the ABLE Exoskeleton). A table with all the results (mean ± standard deviation, % difference between devices, and p-value) from this study can be found in S3 File.

### Safety

A total of 48 AEs (KAFO: n = 31, ABLE: n = 17) were registered during the clinical study, from which only six (KAFO: n = 4, ABLE: n = 2) were reported as device-related by the clinical staff. All the AEs were low severity; and no falls, fractures or device-related SAEs occurred. Other reported events without a medical occurrence were classified as device deficiencies according to ISO 14155:2011. Table 4 shows all the different types of AEs reported during the study classified by device (i.e., KAFO and ABLE) and their cause (i.e., device-related, procedure-related, underlying disease-related, and other causes or undetermined relation).

**Table 4.** Type of adverse events (AEs) that occurred during the clinical investigation.

| Type of AE | Device | Number (% out of total AEs) | Device-related* | Procedure-related* | Underlying disease-related* | Other causes or undetermined relation |
| --- | --- | --- | --- | --- | --- | --- |
| Skin | KAFO | 7 (22.6) | 4 | 2 | 1 | 3 |
|  | ABLE | 7 (41.2) | 1 | 0 | 0 | 6 |
| Pain | KAFO | 18 (58.1) | 0 | 0 | 0 | 18 |
|  | ABLE | 5 (45.5) | 1 | 1 | 0 | 4 |
| Neuropathic pain | KAFO | 4 (12.9) | 0 | 0 | 4 | 0 |
|  | ABLE | 3 (17.7) | 0 | 0 | 3 | 0 |
| Urinary tract | KAFO | 1 (3.2) | 0 | 0 | 1 | 0 |
|  | ABLE | 0 | 0 | 0 | 0 | 0 |
| Spasticity | KAFO | 0 | 0 | 0 | 0 | 0 |
|  | ABLE | 2 (11.8) | 0 | 0 | 2 | 0 |
| Hypotension | KAFO | 1 (3.2) | 0 | 0 | 1 | 0 |
|  | ABLE | 0 | 0 | 0 | 0 | 0 |
| **Total** | **KAFO** | **31 (100)** | **4** | **2** | **7** | **21** |
|  | **ABLE** | **17 (100)** | **2** | **1** | **5** | **10** |

\* Only AEs rated as "Related" are shown. Otherwise, they are classified as "Other causes or undetermined relation". The Safety subsection (Results section) contains a detailed explanation of the latter AEs. In addition, note that one AE can be rated in more than one cause-related.

The most reported type of AE for both devices was pain (KAFO = 18; ABLE = 5). Only one of all the AE of pain (n = 23) was considered to be device-related and occurred when using the ABLE Exoskeleton. This AE consisted of pain in the left hand caused by rhizarthrosis in one of the participants (diagnosed prior to the study), which was intensified by the use of the walker when using the exoskeleton. The issue was resolved by applying a bandage to the affected area of the hand to immobilize the fingers. From the other four AEs of pain reported when walking with the robotic device, two were rated as likely underlying disease-related, which were resolved with resting and thermotherapy, and the other two were low levels of pain in the shoulder that reduced or disappeared during the same training session. Regarding the use of KAFOs, all 18 reported AEs of pain were assigned to other or undetermined causes. Eight of them, classified as likely procedure-related, were from the same participant, the one with rizharthrosis who was very skilled at controlling the KAFOs and asked therapists for more intensive training; causing pain in both hands and arms. To mitigate the pain, additional stretching after the training sessions was applied in the affected areas. From the other 10 AEs of pain issues when walking with KAFOs, four were rated as likely underlying disease-related and affecting the lower back; and the rest were one-time pains without major complications rated as unlikely procedure-related and affecting the arms, neck, and back for causes such as participating in sports the day before the session.

In addition to the aforementioned AEs related to pain, there were two participants with neuropathic pain (one in the dorso-lumbar area and one in the supralesional spinal cord) that experienced pain in all the training sessions and with both devices. However, while one of them kept the same level of pain in the affected area across sessions and devices, the other participant kept the pain constant only when using the robotic device and it increased across sessions when using the KAFOs (2-3 values in the VAS between the beginning and end of the session). This last participant also commented that neuropathic pain decreased during the following days after walking with the ABLE Exoskeleton. In addition, there was a third participant with neuropathic pain in the right lower leg that felt foot pain in almost all the sessions during the training with the ABLE Exoskeleton, and only in one session during KAFOs training (foot and ankle pain). All the AEs for neuropathic pain were rated as underlying disease-related.

The total number of AEs due to skin lesions was lower (KAFO = 7; ABLE = 7), though five of them were considered device-related (KAFO = 4; ABLE = 1). Only one mild pressure injury (grade I according to the EPUAP scale) was reported for the ABLE Exoskeleton and it was due to misuse of the device. The participant wore shorts in one of the training sessions that produced chafing on the skin in contact with the right thigh support. The participant stopped wearing shorts and the skin recovered after two sessions. Contrary to the use of the robotic exoskeleton, device-related skin lesions while using KAFOs were more severe. One of the participants experienced skin irritation in the penis due to friction with the KAFOs during the last evaluation session (i.e., session 10). Moreover, the urine collector intensified the pain and irritation, developing into a little wound. The wound was still under recovery by the end of the study but healing adequately. Another participant suffered a grade II skin damage in both ankles during the second training session with KAFOs. For the following sessions, gauze bandages were used to protect the affected zones and the wounds were completely recovered by the follow-up session. It should be mentioned that this participant started the study with wounds in both ankles (grade I in the right ankle, and grade II in the left ankle) that recovered during the exoskeleton training (their starting device) without additional care and were completely healed before starting with KAFOs. The same happened with two other participants that started the study with wounds in the glutes (grade I) and ankles (grade I), and the right ankle (grade II), respectively. These participants began the training in the ABLE group and the wounds were totally recovered in session 4 (glutes) and session 10 (ankles) with the ABLE Exoskeleton, and session 4 with the KAFO (right ankle), respectively. Finally, another participant experienced a grade I wound on the right ankle due to chafing with the KAFO (their end device) during session 9. The wound was still recovering by the end of the study.

Other AEs with minor complications and not related to the devices were reported during the study. One of the participants reported having been hospitalized over the weekend due to urinary tract infection (UTI) during KAFO training. The participant received treatment and recovered without problems. Another participant experienced spasticity in the abdomen and legs during the first two sessions with the ABLE Exoskeleton. However, after increasing the basal dose of their baclofen regimen, the appearance of spasticity reduced and the participant was able to continue the study protocol without further issues. Finally, one of the participants suffered an isolated episode of orthostatic hypotension at the start of the first evaluation session (i.e., session 5) with the KAFOs, which was classified as likely-related to the underlying disease.

Lastly, two device deficiencies (KAFO: n = 1, ABLE: n = 1) were reported during the clinical investigation. Regarding the robotic device, a wrong length adjustment of both shank and thigh supports that was considered as “use error” made it impossible for the exoskeleton to fully extend the knees, resulting in an excessive motor torque demand that the exoskeleton could not reach. This made both the patient and the therapist feel the knee joint of the device was not exerting enough force. However, the exoskeleton successfully helped the participant to sit down and after a proper size configuration of the device, the issue was solved without complications. With regard to the KAFO deficiency, one of the participant’s KAFOs were not rigid enough at the knee joint, producing a flexion of around five degrees while the participant remained upright. The KAFOs were fixed and the participant could continue the study without problems.

### Feasibility and usability

#### Therapy metrics

The average therapy time per session for all the participants was 54:07 ± 03:37 mm:ss for the ABLE Exoskeleton and 52:34 ± 5:42 mm:ss for the KAFOs (p = 0.383). At the end of the training period (i.e, session 9) with the ABLE Exoskeleton, participants were capable to reach a therapy time of around one hour (60:53 ± 08:38 mm:ss) on average (Table 5), increasing by 10 minutes the time of the first session (50:24 ± 14:07 mm:ss). During this first session, participants walked almost 8 minutes on average (07:54 ± 08:51 mm:ss) and stood for 30 minutes (30:30 ± 13:39 mm:ss). Participants did 242.70 ± 371.46 steps on average, and covered a distance of 83.49 ± 146.75 m at an average gait speed of 0.09 ± 0.07 m/s. By session 9, these values were nearly doubled (walking time: 14:06 ± 7:01 mm:ss; steps: 414.67 ± 334.08 steps; distance: 145.43 ± 134.24 m; gait speed: 0.15 ± 0.07 m/s), except for the standing session that remained constant (30:26 ± 10:00 mm:ss). The *Learning process of using the ABLE Exoskeleton* section examines deeper into these therapy metrics.

**Table 5.** Feasibility and usability metrics.

| | KAFO (Mean ± SD [range]) | ABLE (Mean ± SD [range]) | p-value |
| --- | --- | --- | --- |
| **Therapy Metrics (Session 9)** |  |  |  |
| Therapy time (mm:ss) | 56:30 ± 07:50 [45:00, 70:00] | 60:53 ± 09:10 [47:00, 70:00] | 0.407 |
| Standing time (mm:ss) | - | 30:26 ± 10:00 [19:00, 46:00] | NA |
| Walking time (mm:ss) | - | 14:06 ± 7:01 [6:00, 28:00] | NA |
| Distance (m) | - | 145.43 ± 134.24 [32.21, 461.85] | NA |
| Steps (number) | - | 414.67 ± 334.08 [133, 1250] | NA |
| Average gait speed (m/s) | - | 0.15 ± 0.07 [0.05, 0.26] | NA |
| **Donning/Doffing (Session 10)** |  |  |  |
| Donning time (mm:ss) | 5:05 ± 01:34 [02:39, 07:40] | 04:19 ± 2:06 [01:33, 07:43] | 0.098 |
| Doffing time (mm:ss) | 01:55 ± 00:57 [00:30, 03:28] | 01:58 ± 00:52 [00:31, 03:26] | 1 |
| Donning + Doffing time (mm:ss) | 07:00 ± 02:24 [03:09, 11:08] | 06:17 ± 02:54 [02:04, 11:09] | 0.57 |
| **Clinical outcomes** |  |  |  |
| Distance 6MWT (m) - Session 5 | 35.52 ± 28.20 [12, 98] | 38.86 ± 28.00 [9, 85] | 0.477 |
| Distance 6MWT (m) - Session 10 | 46.30 ± 30.78 [11, 95] | 47.13 ± 29.94 [20, 107] | 0.906 |
| Gait speed 10MWT (m/s) - Session 5 | 0.13 ± 0.09 [0.03, 0.29] | 0.14 ± 0.10 [0.04, 0.32] | 0.557 |
| Gait speed 10MWT (m/s) - Session 10 | 0.17 ± 0.11 [0.04, 0.35] | 0.17 ± 0.10 [0.07, 0.33] | 0.492 |
| Cadence 10MWT (step/min) - Session 5 | 25.65 ± 11.86 [11.72, 44.62] | 23.03 ± 14.69 [10.77, 53.68] | 0.695 |
| Cadence 10MWT (step/min) - Session 10 | 28.91 ± 13.51 [12.78, 48] | 26.62 ± 14.28 [12, 50.53] | 0.695 |
| Time TUG (mm:ss) - Session 5 | 02:05 ± 01:32 [00:43, 05:26] | 02:03 ± 00:54 [00:40, 03:31] | 0.846 |
| Time TUG (mm:ss) - Session 10 | 01:41 ± 01:11 [00:35, 04:36] | 01:42 ± 00:53 [00:37, 03:29] | 0.359 |
| **Therapy activities\*** |  |  |  |
| Sit-to-stand (session) | 1.10 ± 0.32 [1, 2] | 1.70 ± 1.89 [1, 7] | NA |
| Stand-to-sit (session) | 1.80 ± 2.20 [1, 8] | 1.20 ± 0.42 [1, 2] | NA |
| Walking 10 m without stopping (session) | 3.20 ± 2.49 [1, 8] | 3.80 ± 2.15 [1, 8] | NA |
| Turn 180° (session) | 2.00 ± 1.05 [1, 4] | 3.30 ± 1.95 [1, 8] | NA |
| Balance abilities (session) | 1.43 ± 1.45 [1, 3.5] | 1.58 ± 1.09 [1, 3] | NA |
| Walking abilities (session) | 2.34 ± 1.44 [1, 4.6] | 3.14 ± 1.95 [1, 6.6] | NA |
| Advanced abilities (session) | 4.58 ± 2.72 [1.5, 6.9] | 5.75 ± 2.58 [2.8, 8.08] | NA |

\* To report the results of the balance, walking, and advanced abilities, we calculated the average number of sessions that patients required to complete the activities in each ability group. These individual values were used to calculate the mean, standard deviation, and range for these metrics.
NA: not applicable

Regarding the KAFOs, participants were able to increase the duration of the training significantly throughout the study (session 1: 41:15 ± 10:15 mm:ss; session 9: 56:30 ± 07:50 mm:ss; p = 0.033).

#### Donning and doffing

Fig 3 shows the average time and the LOA needed to don and doff the devices in each of the study sessions (i.e., gait training sessions plus evaluation sessions).

**Fig 3.**
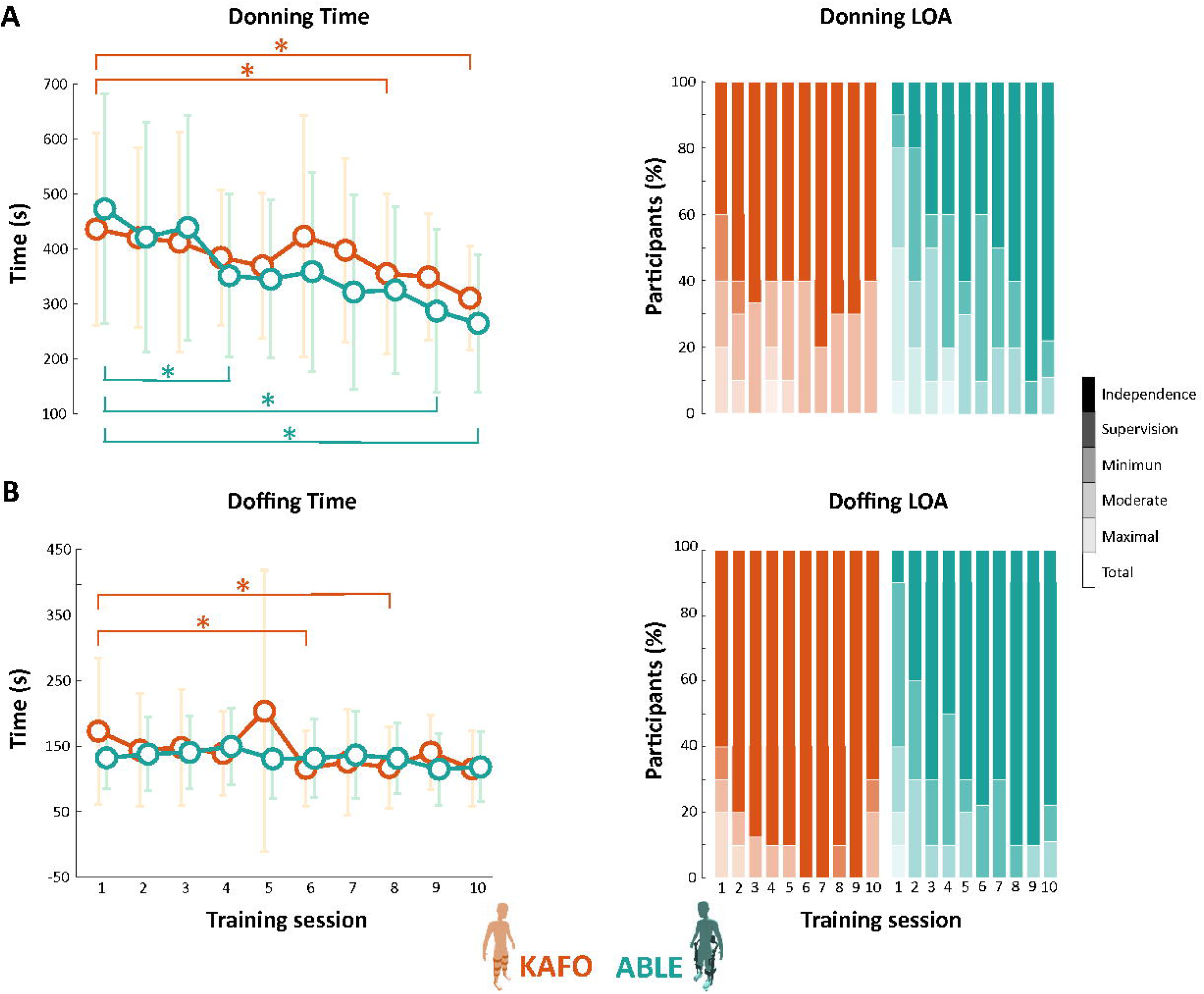
(**a**) Time needed and LOA required for donning both the KAFOs and the ABLE Exoskeleton for all the sessions of the study. (**b**) Time needed and LOA required for doffing both the KAFOs and the ABLE Exoskeleton for all the sessions of the study. Dots indicate the mean value of all the participants. Faded lines show the standard deviation. Stars (*) indicate statistically significant differences (p *<* 0.05).

Both donning and doffing times were very similar between devices across the study. In session 1, donning time with KAFOs was 30 seconds faster than with the ABLE Exoskeleton (KAFO: 07:12 ± 02:56 mm:ss; ABLE: 07:48 ± 03:30 mm:ss; p = 0.432). But this changed in session 4, where donning with ABLE was faster than with KAFOs (KAFO: 06:19 ± 02:04 mm:ss; ABLE: 05:46 ± 02:29 mm:ss; p = 0.432), and this trend was kept until the last session (KAFO: 05:05 ± 01:34 mm:ss; ABLE: 04:19 ± 02:06 mm:ss; p = 0.098). Reduction in donning time from session 1 to 10 was significant for both devices (KAFO: p = 0.024; ABLE: p = 0.027).

Similar to the time needed for donning and doffing, participants experienced a higher improvement (i.e., more independence) in terms of LOA for donning the ABLE Exoskeleton. In session 1, only 50% of the participants could do the donning of the robotic device with minimum assistance or less. However, this improved throughout the training: at the first evaluation session (i.e., session 5) all participants were able to don the device with minimum assistance or less, and at the end of the training almost all the participants (80%) were able to complete donning independently. Previous experience using KAFOs could be observed from the first session, where 80% of the participants did the donning of the KAFOs with minimum assistance or less. At the last session, all the participants managed to perform the donning independently (60%) or with supervision (40%).

#### Step initiation

Already in session 2, participants walked using the automatic step initiation mode for more than half of the session time (average: 62.40 ± 25.48 % of the session time); and from session 7 to session 10, it was practically for the whole session (more than 90% of the session time, on average). In addition, in the second evaluation session (session 10), nine out of the 10 participants completed the standardized clinical tests (i.e., TUG, 10MWT, and 6MWT) with the automatic step initiation mode and used the remote controller to control state transitions themselves.

#### Therapy activities

Table 5 and Fig 4 show the average number of sessions that participants needed to perform each of the therapy activities with minimum assistance or less. When using the ABLE Exoskeleton, all the participants were able to perform *sit-to-stand* and *stand-to-sit* transitions with minimum assistance or less within the first two training sessions (sit-to-stand: 1.70 ± 1.89 sessions; stand-to-sit: 1.20 ± 0.42 sessions); except for one participant, who needed 7 sessions to perform *sit-to-stand* with minimum assistance. Highly similar results were found for the KAFO group, for which all the participants completed both activities within the first two sessions with minimum assistance or less (sit-to-stand: 1.10 ± 0.32 sessions; stand-to-sit: 1.80 ± 2.20 sessions), with the exception of one participant that required 8 training sessions to perform a *stand-to-sit* transition with minimum assistance.

**Fig 4.**
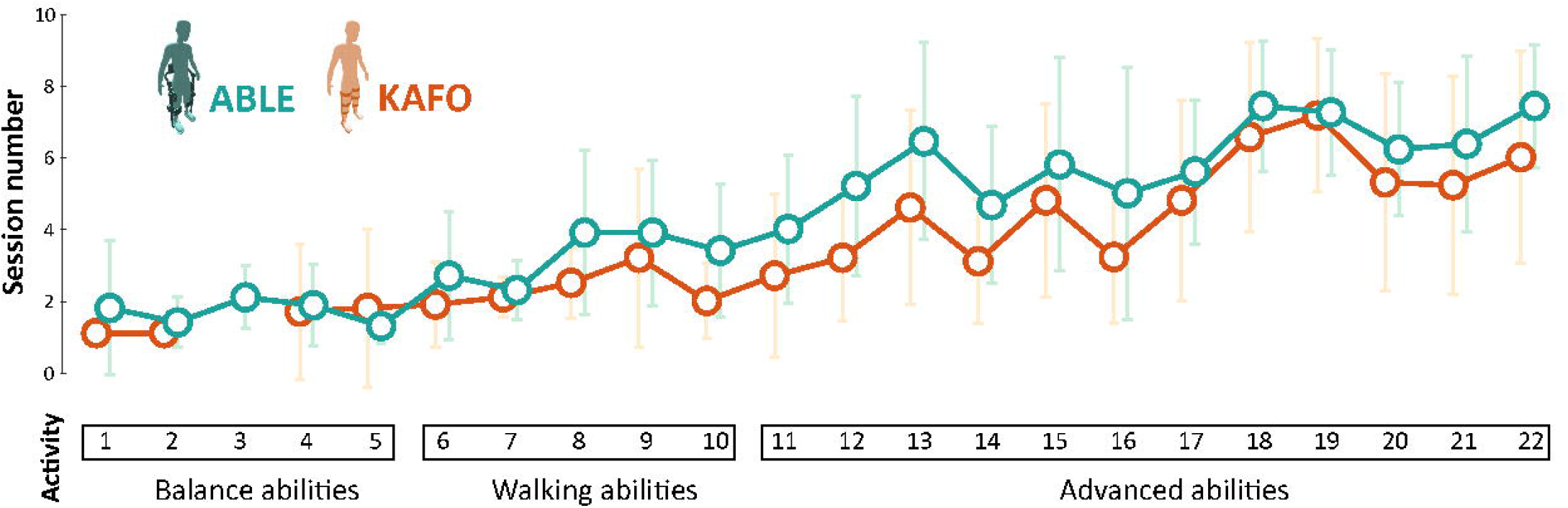
Number of sessions needed to complete each of the therapy activities with minimum assistance or less.

*Walking 10 meters without stopping* with the ABLE Exoskeleton took, on average, almost four sessions (3.80 ± 2.15 sessions), although 70% of the participants did it between sessions 1 and 3. The rest completed the activity in sessions 6 (20%) and 8 (10%). Again, similar results were observed when walking with KAFOs (3.20 ± 2.49 sessions, range = [1, 8]).

Lastly, appreciable differences between devices were noticed in the *turning around* activity (KAFO: 2.00 ± 1.05 sessions, ABLE: 3.30 ± 1.95 sessions). When using the ABLE Exoskeleton, all the participants were able to turn 180º between sessions 1 and 4 with minimum assistance or less, except for one participant that completed the activity in session 8. With KAFOs, only one participant required 4 training sessions to complete the activity, while the rest accomplished it within the first three sessions.

Even having previous experience with the KAFOs, the average number of sessions needed to complete all the balance (KAFO: 1.43 ± 1.45 sessions; ABLE: 1.58 ± 1.09 sessions), walking (KAFO: 2.34 ± 1.44 sessions; ABLE: 3.14 ± 1.95 sessions), and advanced therapy activities (KAFO: 4.58 ± 2.72 sessions; ABLE: 5.75 ± 2.58 sessions) was very similar between devices, and with no more than 1.2 sessions of difference (Table 5 and Fig 4). Note that, despite slight differences, not all the participants completed all the therapy activities with both devices (see S4 Fig); either because they did not accomplish the activity with minimum assistance or less, or because the therapist considered that the patient did not have the skill to perform the activity yet.

#### Clinical outcomes

Fig 5 shows the results for the three standardized clinical tests. On average, participants improved from the first evaluation session (i.e., session 5) to the final evaluation session (i.e., session 10) in all the tests and for both devices. Nonetheless, significant differences, comparing the first and the final evaluation session, were found only for: distance covered during the 6MWT for both KAFO (session 5: 38.86 ± 28.00 m; session 10: 46.30 ± 30.78 m; p = 0.014) and ABLE (session 5: 35.52 ± 28.20 m; session 10: 47.13 ± 29.94 m; p = 0.004); time needed to complete the TUG when using KAFOs (session 5: 02:05 ± 01:32 mm:ss; session 10: 01:41 ± 01:11 m; p = 0.006); gait speed during the 10MWT when walking with KAFOs (session 5: 0.13 ± 0.089 m/s; session 10: 0.17 ± 0.11 m/s; p = 0.004); and cadence during the 10MWT when using KAFOs (session 5: 25.65 ± 11.86 step/min; session 10: 28.91 ± 13.51 step/min; p = 0.014).

**Fig 5.**
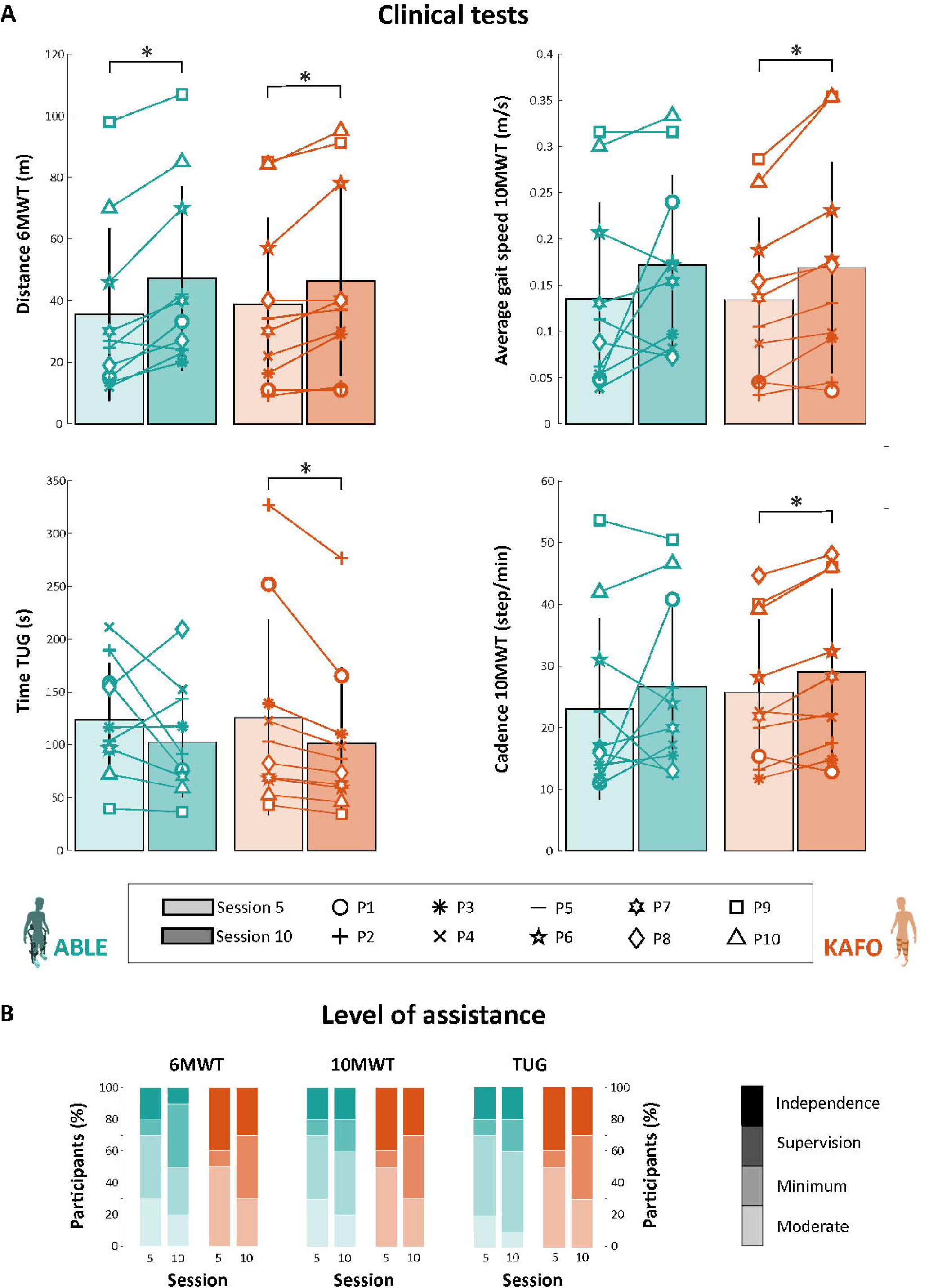
(**a**) Distance covered during the 6MWT, time needed to complete the TUG, gait speed during the 10MWT, and cadence during the 10MWT. Bar plots show the mean and standard deviation from all the participants. (**b**) LOA required to complete the 6MWT, the 10MWT, and the TUG. Stars (*) indicate statistically significant differences (p *<* 0.05).

No significant differences were found when comparing between devices (i.e., KAFO and ABLE) in either session 5 (6MWT: p = 0.477; TUG: p = 0.846; 10MWT gait speed: p = 0.557; 10MWT cadence: p = 0.695) or session 10 (6MWT: p = 0.906; TUG: p = 0.359; 10MWT gait speed: p = 0.492; 10MWT cadence: p = 0.695).

The LOA required to perform the clinical tests was similar between devices. In session 5, 70% of the participants walked with the robotic exoskeleton with minimum assistance or less during the 10MWT and the 6MWT; and 80% during the TUG. Changes occurred in session 10, where 80% of the participants required minimum assistance or less for the 10MWT and the 6MWT, and 90% for the TUG. The rest of the participants needed moderate assistance to perform the tests. When walking with KAFOs, all the participants were able to complete the three tests with minimum assistance or less in both session 5 and session 10. In session 10, only 30% of the participants walked with minimum assistance, and the others completed the tests with independence (30%) or supervision (40%).

### Learning process of using the ABLE Exoskeleton

Note that for this section, P9 and P10 were not included in the statistical analysis, since they were identified as outliers in nearly all the sessions for all the therapy metrics recorded with the ABLE Exoskeleton. Also, note that the evaluation sessions were not considered in the statistical analysis as they had notable differences with respect to the gait training sessions. Therefore, only sessions 1 to 4 and 6 to 9 were considered in this regard.

#### Therapy time

The mean therapy time was very constant across sessions (Fig 6), keeping an average time per session between 48 and 60 minutes (53:36 ± 12:06 mm:ss). The 53.79% of this time was spent standing (28:42 ± 9:36 mm:ss). However, participants gradually increased their walking time over the training sessions, showing a significant difference between the first and the final training session (session 1: 03:52 ± 02:51 mm:ss; session 9: 12:22 ± 05:42 mm:ss; p = 0.008). The walking-standing time ratio increased almost threefold from the first to the last session (session 1: 14.66 ± 9.40 %; session 9: 41.82 ± 10.96 %; p = 0.008).

**Fig 6.**
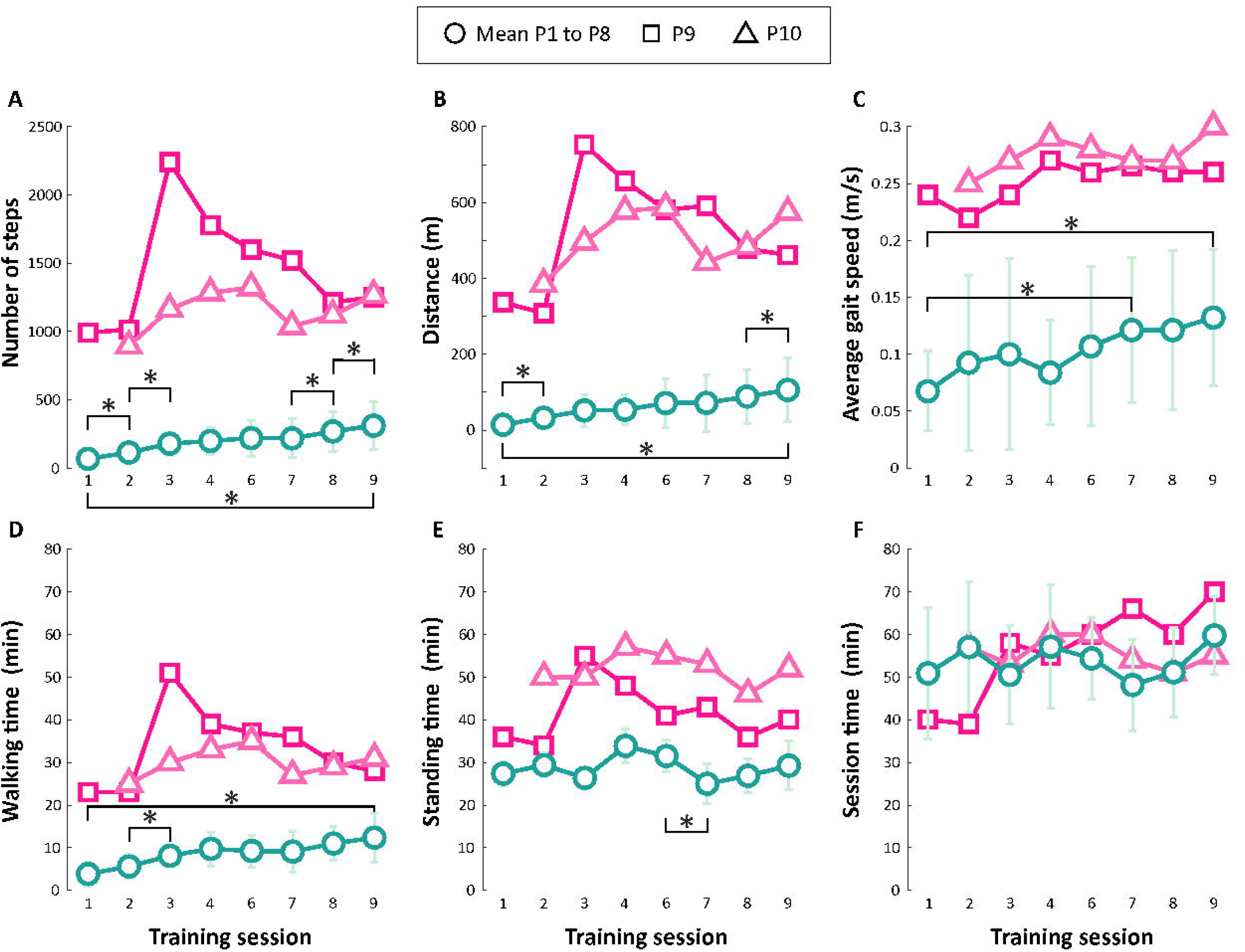
Mean and standard deviation for all the therapy metrics recorded by the ABLE Exoskeleton during all the training sessions (i.e., sessions 1 to 4 and 5 to 9): (**a**) Number of steps, (**b**) distance, (**c**) average gait speed, (**d**) walking time, (**e**) standing time, and (**f**) training session time. Stars (*) indicate statistically significant differences (p *<* 0.05).

Despite the similarities in the therapy time, P9 and P10 stood out for a higher standing time (average: 84.41 ± 13.47 % of the therapy time) with a substantially higher walking-standing time ratio (P9 average: 79.10 ± 9.99 %; P10 average: 57.88 ± 5.04 %).

#### Number of steps

The mean number of steps (Fig 6) significantly increased by 4.57 times from session 1 to 9 (session 1: 67.86 ± 45.49 steps; session 9: 310.25 ± 177.08 steps; p = 0.008). Participants had a rapid and significant improvement from session 1 to 2 (session 1: 67.86 ± 45.49 steps; session 2: 113.63 ± 55.21 steps; p = 0.030), and from session 2 to 3 (session 2: 113.63 ± 55.21 steps; session 3: 178.38 ± 84.9234 steps; p = 0.023). After that, they kept relatively constant for three sessions until a new significant increment in the number of steps by the end of the training period, from sessions 7 to 8 (session 7: 217.50 ± 141.17 steps; session 8: 268.25 ± 145.94 steps; p = 0.039) and 8 to 9 (session 8: 268.25 ± 145.94 steps; session 9: 310.25 ± 177.08 steps; p = 0.050).

P9 and P10 did, on average, 1451.50 ± 396.65 steps and 1155.00 ± 141.68 steps per session, respectively. Although they increased the number of steps from session 1 to 9, they did not follow a progressive rise. P9 was able to walk up to 2242 steps in one single session (session 3).

#### Distance

Highly correlated with the number of steps, the distance walked during the training sessions showed a similar trend (Fig 6). Participants had a significant increase at the beginning of the training period, from session 1 to 2 (session 1: 14.20 ± 9.09 m; session 2: 32.50 ± 24.48 m; p = 0.016); and at the end of the training, from session 8 to 9 (session 8: 88.35 ± 70.72 m; session 9: 105.88 ± 84.13 m; p = 0.016). From session 1 to 9, the distance covered significantly increased by 7.56 times (session 1: 14.20 ± 9.09 m; session 9: 105.88 ± 84.13 m; p = 0.008).

P9 and P10 were again superior to the other participants. The mean distance walked was 520.71 ± 143.78 m and 506.47 ± 71.37 m, respectively; with a maximum distance of 752.94 m for P9 (session 3) and 587.07 m for P10 (session 6).

#### Average gait speed

Average gait speed increased gradually across the training sessions (Fig 6). However, it was not until session 7 that participants significantly walked faster than the initial session (session 1: 0.07 ± 0.04 m/s; session 7: 0.12 ± 0.06 m/s; p = 0.008). From that session, average gait speed continued increasing until the end of the training with the ABLE Exoskeleton (session 9: 0.13 ± 0.06 m/s).

P9 and P10 also increased their gait speed over the sessions, but with more fluctuations than the rest of the participants. They finished the training with an average gait speed of 0.29 m/s and 0.32 m/s, respectively.

## Discussion

This randomized, crossover clinical trial is, to the best of our knowledge the first one that compares the use of a knee-powered lower limb exoskeleton against conventional KAFOs in individuals with motor-complete SCI in terms of safety, feasibility, and usability. Our findings suggest that the ABLE Exoskeleton is as feasible and simple to use as traditional KAFOs, and better in terms of safety.

### Safety

Understanding better the AEs and risks of using wearable exoskeletons through comprehensive reporting is needed to develop safe devices [6, 49]. To this end, we performed a detailed and extensive safety assessment of the ABLE Exoskeleton throughout the clinical trial. The same assessment was done for the KAFOs to be able to carry out a comparison in terms of safety between conventional mechanical and new robotic devices for overground gait training.

In this study, a total of 99 and 100 gait training sessions were conducted with the ABLE Exoskeleton and with KAFOs, respectively. A total of 17 AEs were reported when using the robotic device (see Table 4), from which only two were rated as device-related due to skin lesion and pain. On the other hand, 31 AEs were recorded when walking with KAFOs, four of them rated as device-related and due to skin lesions. Additionally, one device deficiency was reported for each device. In both cases, the issue was resolved without any negative consequences for the participants.

Skin damage in areas that are in contact with the device is the most common risk of using wearable exoskeletons [6, 24, 50], even in those with FDA and CE approval: Rewalk [18], Indego [20], and Ekso [3, 17]. However, only one low-severity AE related to skin damage was reported when walking with the ABLE Exoskeleton due to skin redness. This might indicate that the device was able to adjust well to all participants. Moreover, user-exoskeleton contact areas are not in high-risk zones for skin damage.

The same cannot be said for the KAFOs, where four out of the seven AEs of skin type were related to the device and had higher severity. Despite being personalized to the anatomy of the leg, the majority of the KAFOs’ thermoplastic surface is directly in contact with the participant’s legs, which increases the risk of suffering skin wounds, especially in areas that are in contact with bones (e.g., the ankles). Therefore, in that sense, wearable exoskeletons, like the ABLE Exoskeleton, that greatly reduce direct contact with the participant’s body through gel padding might significantly reduce the risk of skin damage.

Pain is another well-known potential risk when using robotic exoskeletons [3, 6, 22, 24, 34]. In this study, only one AE (out of five) was classified as device-related for the ABLE Exoskeleton and it was in the left hand of one of the participants. The other reported pains (n = 4) were sporadic pains in the shoulder, arms, or back, that participants felt before the session and remained at the same level of pain (according to the VAS) or even decreased after the training session. This suggests that using the ABLE Exoskeleton not only does not produce pain, but in case there is an existent pain before its use, this one does not increase and could even be reduced. Regarding the pain levels when using the KAFOs, half of the reported AEs (9 out of 18) were one-time pains that lasted no more than two consecutive sessions and were mainly located in the lower back. The other half were concentrated on one participant who mastered the use of KAFOs and asked for very intensive training, which produced pain in the arms and hands after almost each training session. Pain in these aforementioned areas is common when using KAFOs due to atypical gait patterns (e.g., circumduction and hip hiking [9, 33, 51–54]) and excessive use of the walker to maintain balance while walking [55, 56]. This noticeable difference in the number of AEs of pain between devices (KAFO = 18; ABLE = 5) may be due to the more physiological gait pattern provided by the ABLE Exoskeleton (topic previously addressed in [33]), which allows users to rely less on walking aids.

Finally, a few underlying-disease-related AEs occurred during the study, as expected due to the proneness of these issues in people with SCI, which have also been reported in other exoskeleton trials [21, 22].

All of these findings show that the ABLE Exoskeleton is not only comparable to the KAFOs in terms of safety (or even better), but also to other commercially available exoskeletons. Yet, comparing the safety among robotic exoskeletons is challenging since studies tend to omit relevant details and, sometimes, do not explicitly report whether adverse events occurred [49].

### Feasibility and usability

In general, participants stood up for at least 30 minutes in all the training sessions with the ABLE Exoskeleton, meeting the recommendations for people with SCI drawn by Paleg & Livingstone in their systematic review [57]. The walking time ended up representing half of the standing time (44.95 ± 13.61 %) by the end of the training and increased by 78.61% from the first to the last training session. Moreover, participants required about five sessions to control the automatic step initiation mode (i.e., users can initiate steps on their own), and from session 7 on (i.e., session 7 to 10), the majority of the therapy time was performed with the automatic step initiation mode (above 90% of the therapy time, on average). These results show the participants’ high adaptability to the device, which is crucial for delivering intensive training from the very beginning of the rehabilitation process.

Over the course of the study, participants improved in all gait parameters. Participants increased the average distance covered (74%), the number of steps (71%), and the gait speed (50%) along the eight training sessions. Unfortunately, these metrics cannot be compared with the KAFOs since this type of data was not collected. Likewise, due to the lack of information provided in similar studies and the divergence in the clinical protocols and/or population, direct comparisons with other exoskeletons are difficult to be done [3, 17, 21, 22]. The feasibility and usability of the ABLE Exoskeleton were also assessed in [24]. The latter study reported improvements of 290%, 300%, and 180% in the distance covered, the number of steps, and the gait speed, respectively; which are higher than the ones found in our work. Such variation can be explained by the fact that the LOI (from C5 to L3), the severity of the injury (AIS A to D), and the time since injury (mainly subacute: *<*1 year after injury) of the studied population in [24] differed from the ones in our study. This hinders the direct comparison of gait parameters’ improvement between studies. Nevertheless, this comparison reveals how population-reliant wearable exoskeletons can be.

The amount of assistance and time required to perform both the donning and doffing of the device decreased through the training program. Almost all the participants were able to independently don (n = 8/10) and doff (n = 8/10) the ABLE Exoskeleton after 10 sessions (see Fig 3). The mean average time to don and doff the device in the last session was 06:17 mm:ss, where doffing occurred to be considerably easier (half of the time) for participants than donning. All the participants decreased the time and the LOA for donning and doffing the device during the training period. All of these findings demonstrate an improvement over the donning and doffing times for commercially available exoskeletons (i.e., ReWalk, Ekso, and Indego exoskeletons) used in the clinical setting that have been reported in the literature, where average donning and doffing times were in the range of 05:00 to 18:00 mm:ss and 02:44 to 05:00 mm:ss, respectively, with a LOA from moderate assistance to independence [17, 20, 34, 39]. Compared to the KAFOs, the ABLE Exoskeleton did not show significant differences either for the time or the LOA to don/doff the device. As expected, participants showed better results at the beginning of the study to don/doff the KAFOs than the robotic device, mainly due to previous experience using KAFOs. However, this difference disappeared halfway through the study and even reversed by the end of the study, where the ABLE Exoskeleton stood out for a faster donning time. This result highlights the simplicity to don/doff the ABLE Exoskeleton and its fast learning process, which is supported by [24] in which the average time to don and doff the ABLE Exoskeleton in the last session was 06:50 mm:ss in a population of predominantly acute and subacute SCI individuals (20 out of 24). It is worth noting that the complexity to don and doff robotic exoskeletons is one of the reasons for not adopting them in the clinical setting [58]. However, our results indicate that this should not be a barrier because donning and doffing the ABLE Exoskeleton took about the same amount of time as KAFOs, which are widely accepted.

Nearly all the participants completed all the therapy activities (see Table 2) during the training period, both when using the KAFOs and the ABLE Exoskeleton. Although participants had previous experience with the KAFOs, the number of sessions needed to complete each activity was very similar between devices and it only took generally one session more to achieve the activities using the wearable exoskeleton compared to the KAFOs (see Fig 4). Overall, and for both devices, participants needed, on average, 1 to 2 sessions to perform sit-to-stand and stand-to-sit transitions with minimum assistance or less; and only 3 to 4 sessions to walk 10 meters (without stops) and turn around. In general, by the end of the training, all participants were able to perform around 22 activities of variant difficulty with minimum assistance or less. These data imply that using the ABLE Exoskeleton could be very intuitive, as participants learned to perform the therapy activities almost as quickly as with the KAFOs. These findings also constitute an improvement with respect to other commercially available exoskeletons that reported the LOA to stand, walk, and/or sit with the devices. In the study of Kozlowsky et al. [34], participants (mostly in the subacute phase, 43% with motor incomplete SCI) needed a median of 8 sessions to both walk and stand/sit with minimum assistance or less while using a wearable lower limb exoskeleton. In the study of Gagnon et al. [22], where the population examined was more similar to that of this study (mostly chronic with motor complete SCI), 15 sessions (plus two familiarization sessions) were needed to see all the participants walking with minimum assistance or less; and all of them needed moderate to maximal assistance for all sit-to-stand and stand-to-sit transitions throughout the training sessions.

Overall, all the participants became more skilled over time with both devices, showing improvements in the three standardized clinical tests (i.e., 10MWT, 6MWT, and TUG) (see Fig 5). In regards to the ABLE Exoskeleton, significant improvements from the first to the final evaluation session (i.e., sessions 5 and 10, respectively) were only found for the distance covered during the 6MWT. However, the minimal clinically important difference (MCID) was exceeded for the TUG (MCID: 10.8 s; ABLE difference: 21.15 s), according to the values reported by Lam et al. in their systematic review [59]. Possibly, the large variability noted among participants and the limited number of participants reduced the statistical power to get significant differences in the TUG test. During KAFOs training, the MCID was also achieved for the TUG (KAFO difference: 24.55 s), and all the metrics measured during the clinical tests presented significant differences between the two evaluation sessions. These results reveal that despite having previous experience walking with KAFOs (being most of the participants regular users), they still had room for improvement. Therefore, this might indicate that 8 training sessions were not enough to see the complete learning process of the ABLE Exoskeleton and, moreover, that further and significant improvements could appear with more extensive training. Despite that, noticeable improvements in the clinical tests when using the ABLE Exoskeleton were achieved at the same time that participants reduced the necessary LOA to perform them, where roughly all the participants completed the tests with minimum assistance or less (n = 9/10 for the TUG; n = 8/10 for the 10MWT and 6MWT). The LOA for using the KAFOs also improved between the evaluation sessions but on a smaller scale, since participants showed high independence using the passive orthoses from the very beginning of the study due to their prior experience.

In the final evaluation session, participants walked with the ABLE Exoskeleton a total distance of 47.13 ± 29.94 m (range = [20, 107]), completed the TUG in 02:03 ± 00:54 mm:ss (range = [00:40, 03:31]), and walked the 10MWT with a gait speed of 0.17 ± 0.10 m/s (range = [0.07, 0.33]) and a cadence of 23.03 ± 14.69 steps/min (range = [10.77, 53.68]). Very similar values were measured in the final evaluation session with KAFOs, showing no significant differences between devices either for the final or the first evaluation session. These values are, on average, distant from the values reported for other commercially available wearable exoskeletons for overground walking [60, 61]. However, this comparison should be taken with caution, considering that the exoskeletons from the aforementioned studies offered higher walking assistance through hip and knee actuators, the LOI of the studied population differed from the current study (e.g., includes motor incomplete paraplegia), and participants trained an average of 21 ± 13 sessions. Further clinical studies with a higher number of sessions using the ABLE Exoskeleton are needed for proper comparison to other robotic devices in terms of clinical test performance.

### Learning process

We have seen that donning and doffing the robotic device was relatively easy to learn, especially the doffing process (see Fig 3). By session 2, all the participants already managed to doff the device with minimum assistance or less in a time of 02:18 mm:ss, which is notably faster compared to other commercially available exoskeletons (range: 02:44 to 05:00 mm:ss [17, 20, 34, 39]). This time was kept practically constant during the study with an average reduction of 23 seconds between the first and last session. One reason for this trend may be that the ease and simplicity of doffing the ABLE Exoskeleton allowed participants to quickly learn how to do it at the start of the study, leaving a low margin for improvement. On the other hand, the donning process took more sessions to be learned. It was in sessions 4 and 5 where a substantial reduction in time was observed, being session 5 the time point when all participants needed minimum assistance or less to don the device. From that point onward, participants refined their performance and still showed improvements in the time needed to don the exoskeleton. Particularly, the last two sessions showed a clear improvement in the donning time.

Another argument to consider the ease of use of the ABLE Exoskeleton is the rapidity with which participants completed the therapy activities (see Fig 4 and Table 5). It took, on average, no more than 4 sessions for the participants to operate the device during activities such as sit-to-stand, stand-to-sit, walking 10 meters, and turning around (fundamental activities to control a robotic exoskeleton independently), with minimum assistance or less. Furthermore, almost all the participants were able to perform more challenging activities like *stop in front of a door, open it (inward) and continue walking* (n = 8/10) or *stop near a wall, turn, and lean against it* (n = 9/10) in just 7.13 ± 2.03 and 6.11 ± 1.90 sessions, respectively. Conversely, in the study of [24], participants required more sessions and higher LOA than in the current study to perform therapy activities like walking and sit-stand transitions. An explanation for that may be the difference between acute/subacute individuals with chronic individuals, where the latter are already rehabilitated, used to the injury, more confident and skilled, and, in general, experienced with gait assistive devices. This comparison points out again the relation between exoskeleton performance and population.

For the therapy metrics, we detected two participants that performed unarguably better than the rest (see Fig 6). These participants had both a very active lifestyle, were very fit, and had a low LOI (T11 and T10), which are factors that have been identified as the main performance predictors in people with SCI when learning to use a wearable exoskeleton [37]. Moreover, they had previous experience with other lower-limb exoskeletons. These two participants were able to stay more time standing and walking than the rest, giving them more time to practice and learn. Likewise, they showed better results than the other participants in the 6MWT and the 10MWT (see Fig 5). However, in the TUG and during the therapy activities, the differences were not so pronounced. Focusing on the other participants, we clearly observed a progressive improvement during the training sessions in terms of the number of steps, distance walked, gait speed, and walking time. Yet, it seems that participants did not reach a plateau by the end of the study. This is more notorious, for example, in the number of steps and distance where still significant improvements occurred between consecutive sessions at the end of the training period. This slow but constant tendency certainly shows that participants still had room for improvement, and more sessions would be needed to reach their optimal performance.

Generally speaking, the ABLE Exoskeleton seems to be an easy-to-use wearable exoskeleton that people with complete SCI can learn to operate with high independence in approximately 5 training sessions. In fact, despite having previous experience with KAFOs use, participants quickly mastered the same skills with the ABLE Exoskeleton and showed equivalent results in the clinical tests (i.e., 10MWT, 6MWT, and TUG).

## Limitations and future work

There were some limitations in the present study. While having a diverse range of injury levels among study participants to evaluate the assistive devices is interesting, this diversity may have added more variability to the data, limiting the statistical significance of the results. Another limitation is that the participants were already used to walking with KAFOs, which may have distorted the comparison. However, comparing subjects who are new to KAFOs and new to the ABLE Exoskeleton is complicated since KAFOs are usually prescribed by the hospital once the patient is discharged at home. Also, the inability to measure therapy metrics such as the number of steps or standing/walking time when using the KAFOs limited the comparison between conventional passive orthoses and robotic gait therapy during training. In like manner, measuring the progress of the LOA needed to complete the therapy activities, instead of only the first session when the activity was completed, could have given a better understanding of the learning process to perform exoskeleton skills. In consonance with that, the method used to evaluate the therapy activities’ progression may have confounded its analysis, as the participants were not required to follow the established order, despite the fact that the difficulty of the activities gradually increased. Furthermore, we have seen that a larger number of training sessions would be needed to capture the complete learning process of using the ABLE Exoskeleton. Finally, taking into account the low number of sessions that participants needed to learn to control the ABLE Exoskeleton, further studies will conduct the baseline session earlier in the training period to obtain more realistic baseline values.

## Conclusion

The results of this clinical study showed that the ABLE Exoskeleton is safe, feasible, and easy to use for gait training in people with motor complete SCI (AIS A and B) with neurological levels ranging from T4 to T12 in a rehabilitation hospital setting. Only 17 low-severe AEs were reported for the robotic device during the study, being only 2 of them device-related. Donning and doffing the device was feasible for all the participants, which needed minimum assistance or less. Overall, participants rapidly improved their skills with the exoskeleton throughout the training, demonstrating improvements in therapy metrics, progressing with the therapy activities, and accomplishing the most challenging abilities. Furthermore, the present study was the first to compare the use of KAFOs (standard of care) against a wearable lower limb exoskeleton (i.e., the ABLE Exoskeleton) for assisting gait in people with SCI in terms of safety, feasibility, and usability. The exoskeleton proved to be superior to KAFOs with regard to safety and demonstrated to be as practical and easy to use as the conventional passive orthoses. Finally, the insights gained from this clinical trial have been critical in helping the engineers at ABLE Human Motion develop the next version of the ABLE Exoskeleton that includes powered actuation in the hip joints. Clinical trials will be conducted in the future using the new design of the ABLE Exoskeleton.

## Data Availability

The data underlying the results presented in the study are available from ABLE Human Motion, Barcelona, 08028, Spain (https://www.ablehumanmotion.com/).

## Acknowledgements

The authors thank Anna Mas, Miller Espinosa, Vicent Pérez, and Iris Bálcazar for their help and support during the study. Furthermore, the authors would like to thank Pablo Peret, Marc Mesia, and Marc Serra for their help during the training, and Joan C. Martori for his help with the statistical analysis. Finally, the authors thank Katlin Kreamer-Tonin for proofreading the final version of the manuscript.

## Supporting information

**S1 File. Clinical Research Ethics Committee’s resolution**.

**S2 Table. Inclusion/exclusion criteria**.

**S3 File. Detailed results**. Data sheet with all the results presented in this manuscript.

**S4 Fig. Number of participants that completed the ability activities**. Number of participants that completed each of the ability activities with no more than minimum assistance for both devices.

**S5 File. Trial study protocol**.

## References

1. World Health Organization (WHO). International perspectives on spinal cord injury; Date accessed: 2022-09-15. Available from: https://www.who.int/publications/i/item/international-perspectives-on-spinal-cord-injury.

2. Baunsgaard CB, Nissen UV, Brust AK, Frotzler A, Ribeill C, Kalke YB, et al. Gait training after spinal cord injury: safety, feasibility and gait function following 8 weeks of training with the exoskeletons from Ekso Bionics. Spinal Cord. 2018;56:106–116. doi:10.1038/s41393-017-0013-7.

3. McIntosh K, Charbonneau R, Bensaada Y, Bhatiya U, Ho C. The Safety and Feasibility of Exoskeletal-Assisted Walking in Acute Rehabilitation After Spinal Cord Injury. Archives of Physical Medicine and Rehabilitation. 2020;101(1):113–120. doi:https://doi.org/10.1016/j.apmr.2019.09.005.

4. Eng JJ, Levins SM, Townson AF, Mah-Jones D, Bremner J, Huston G. Use of prolonged standing for individuals with spinal cord injuries. Physical therapy. 2001;81 8:1392–9. doi:10.1093/PTJ/81.8.1392.

5. Sale P, Russo EF, Russo M, Masiero S, Piccione F, Calabró RS, et al. Effects on mobility training and de-adaptations in subjects with Spinal Cord Injury due to a Wearable Robot: a preliminary report. BMC Neurology. 2016;16. doi:10.1186/s12883-016-0536-0.

6. Rodríguez-Fernández A, Lobo-Prat J, Font-Llagunes JM. Systematic review on wearable lower-limb exoskeletons for gait training in neuromuscular impairments. Journal of NeuroEngineering and Rehabilitation. 2020;18. doi:10.1186/s12984-021-00815-5.

7. Merkel KD, Miller NE, Merritt JL. Energy expenditure in patients with low-, mid-, or high-thoracic paraplegia using Scott-Craig knee-ankle-foot orthoses. Mayo Clinic proceedings. 1985;60 3:165–8. doi:10.1016/S0025-6196(12)60213-4.

8. Lavis TD, Codamon L. Lower Limb Orthoses for Persons With Spinal Cord Injury. Atlas of Orthoses and Assistive Devices. 2019;doi:10.1016/B978-0-323-48323-0.00023-8.

9. Harvey LA. Chapter 6 – Standing and walking with lower limb paralysis. In: Management of Spinal Cord Injuries: A guide for Physiotherapists; 2008. p. 107–136.

10. Lawrason S, Tomasone J, Olsen K, Ginis KM. I’m glad I can walk, but sometimes it’s so challenging that it’s an inconvenience to myself and others: physical activity experiences among individuals with spinal cord injury who ambulate. Qualitative Research in Sport, Exercise and Health. 2022;14(6):987–1004. doi:10.1080/2159676X.2022.2046630.

11. Chen G, Chan CK, Guo Z, Yu H. A review of lower extremity assistive robotic exoskeletons in rehabilitation therapy. Critical reviews in biomedical engineering. 2013;41 4-5:343–63. doi:10.1615/CRITREVBIOMEDENG.2014010453.

12. Díaz I, Gil JJ, Sánchez E. Lower-Limb Robotic Rehabilitation: Literature Review and Challenges. J Robotics. 2011;2011:759764:1–759764:11. doi:10.1155/2011/759764.

13. Reinkensmeyer D, Dietz V. Neurorehabilitation technology. 2nd ed. Springer International Publishing; 2016.

14. Esquenazi A, Talaty M. Robotics for Lower Limb Rehabilitation. Physical medicine and rehabilitation clinics of North America. 2019;30 2:385–397. doi:10.1016/J.PMR.2018.12.012.

15. Colombo G, Joerg M, Schreier R, Dietz V. Treadmill training of paraplegic patients using a robotic orthosis. Journal of rehabilitation research and development. 2000;37 6:693–700.

16. Gassert R, Dietz V. Rehabilitation robots for the treatment of sensorimotor deficits: a neurophysiological perspective. Journal of NeuroEngineering and Rehabilitation. 2018;15. doi:10.1186/s12984-018-0383-x.

17. Kolakowsky-Hayner SA, Crew JD, Moran S, Shah A. Safety and Feasibility of using the EksoTM Bionic Exoskeleton to Aid Ambulation after Spinal Cord Injury. Journal of Spine. 2013;2013:1–8. doi:10.4172/2165-7939.S4-003.

18. Yang A, Asselin PK, Knezevic S, Kornfeld SD, Spungen AM. Assessment of In-Hospital Walking Velocity and Level of Assistance in a Powered Exoskeleton in Persons with Spinal Cord Injury. Topics in spinal cord injury rehabilitation. 2015;21 2:100–9. doi:10.1310/sci2102-100.

19. Miller LE, Zimmermann AK, Herbert WG. Clinical effectiveness and safety of powered exoskeleton-assisted walking in patients with spinal cord injury: systematic review with meta-analysis. Medical Devices (Auckland, NZ). 2016;9:455 – 466. doi:10.2147/MDER.S103102.

20. Tefertiller C, Hays K, Jones J, Jayaraman A, Hartigan C, Bushnik T, et al. Initial Outcomes from a Multicenter Study Utilizing the Indego Powered Exoskeleton in Spinal Cord Injury. Topics in spinal cord injury rehabilitation. 2018;24 1:78–85. doi:10.1310/sci17-00014.

21. Baunsgaard CB, Nissen UV, Brust AK, Frotzler A, Ribeill C, Kalke YB, et al. Exoskeleton gait training after spinal cord injury: An exploratory study on secondary health conditions. Journal of rehabilitation medicine. 2018;50 9:806–813. doi:10.2340/16501977-2372.

22. Gagnon DH, Escalona MJ, Vermette M, Carvalho LP, Karelis AD, Duclos C, et al. Locomotor training using an overground robotic exoskeleton in long-term manual wheelchair users with a chronic spinal cord injury living in the community: Lessons learned from a feasibility study in terms of recruitment, attendance, learnability, performance and safety. Journal of NeuroEngineering and Rehabilitation. 2018;15. doi:10.1186/s12984-018-0354-2.

23. Xiang XN, Ding M, yan Zong H, Liu Y, Cheng H, He CQ, et al. The safety and feasibility of a new rehabilitation robotic exoskeleton for assisting individuals with lower extremity motor complete lesions following spinal cord injury (SCI): an observational study. Spinal Cord. 2020;58:787–794. doi:10.1038/s41393-020-0423-9.

24. Wright MA, Herzog F, Mas-Vinyals A, Carnicero-Carmona A, Lobo-Prat J, Hensel C, et al. Multicentric investigation on the safety, feasibility and usability of the ABLE lower-limb robotic exoskeleton for individuals with spinal cord injury: a framework towards the standardisation of clinical evaluations. Journal of NeuroEngineering and Rehabilitation. 2023, (under review);.

25. ReWalk™ Personal Exoskeleton System Cleared by FDA for Home Use; Date accessed: 2023-01-18. Available from: https://ir.rewalk.com/news-releases/news-release-details/rewalktm-personal-exoskeleton-system-cleared-fda-home-use.

26. Indego FDA Clearance for Clinical and Personal Use; Date accessed: 2022-11-30. Available from: https://www.indego.com/indego/us/en/home.

27. Arazpour M, Bani MA, Hutchins SW, Jones RK. The physiological cost index of walking with mechanical and powered gait orthosis in patients with spinal cord injury. Spinal Cord. 2013;51:356–359. doi:10.1038/sc.2012.162.

28. Farris RJ, Quintero HA, Murray SA, Ha KH, Hartigan C, Goldfarb M. A Preliminary Assessment of Legged Mobility Provided by a Lower Limb Exoskeleton for Persons With Paraplegia. IEEE Transactions on Neural Systems and Rehabilitation Engineering. 2014;22:482–490. doi:10.1109/TNSRE.2013.2268320.

29. Yatsuya K, Hirano S, Saitoh E, Tanabe S, Tanaka H, Eguchi M, et al. Comparison of energy efficiency between Wearable Power-Assist Locomotor (WPAL) and two types of knee-ankle-foot orthoses with a medial single hip joint (MSH-KAFO). The Journal of Spinal Cord Medicine. 2018;41:48 –54. doi:10.1080/10790268.2016.1226701.

30. Wu CH, Mao HF, Hu J, Wang TY, Tsai YJ, Hsu WL. The effects of gait training using powered lower limb exoskeleton robot on individuals with complete spinal cord injury. Journal of NeuroEngineering and Rehabilitation. 2018;15. doi:10.1186/s12984-018-0355-1.

31. Kwon SH, Lee BS, Lee HJ, Kim EJ, Lee JA, Yang SP, et al. Energy Efficiency and Patient Satisfaction of Gait With Knee-Ankle-Foot Orthosis and Robot (ReWalk)-Assisted Gait in Patients With Spinal Cord Injury. Annals of Rehabilitation Medicine. 2020;44:131 – 141. doi:10.5535/arm.2020.44.2.131.

32. Chen S, Wang Z, Li Y, Tang J, Wang X, Huang L, et al. Safety and Feasibility of a Novel Exoskeleton for Locomotor Rehabilitation of Subjects With Spinal Cord Injury: A Prospective, Multi-Center, and Cross-Over Clinical Trial. Frontiers in Neurorobotics. 2022;16. doi:10.3389/fnbot.2022.848443.

33. Rodríguez-Fernández A, Lobo-Prat J, Tarragó R, Chaverri D, Iglesias X, Guirao-Cano L, et al. Comparing walking with knee-ankle-foot orthoses and a knee-powered exoskeleton after spinal cord injury: a randomized, crossover clinical trial. Scientific Reports. 2022;12(19150). doi:https://doi.org/10.1038/s41598-022-23556-4.

34. Kozlowski AJ, Bryce TN, Dijkers MP. Time and Effort Required by Persons with Spinal Cord Injury to Learn to Use a Powered Exoskeleton for Assisted Walking. Topics in spinal cord injury rehabilitation. 2015;21 2:110–21. doi:10.1310/sci2102-110.

35. Platz T, Gillner A, Borgwaldt N, Kroll S, Roschka S. Device-Training for Individuals with Thoracic and Lumbar Spinal Cord Injury Using a Powered Exoskeleton for Technically Assisted Mobility: Achievements and User Satisfaction. BioMed Research International. 2016;2016. doi:10.1155/2016/8459018.

36. van Dijsseldonk RB, Rijken H, van Nes IJW, van de Meent H, Keijsers NLW. A Framework for Measuring the Progress in Exoskeleton Skills in People with Complete Spinal Cord Injury. Frontiers in Neuroscience. 2017;11. doi:10.3389/fnins.2017.00699.

37. van Dijsseldonk RB, Rijken H, van Nes IJW, van de Meent H, Keijsers NLW. Predictors of exoskeleton motor learning in spinal cord injured patients. Disability and Rehabilitation. 2019;43:1982 – 1988. doi:10.1080/09638288.2019.1689578.

38. Spungen AM, Asselin PK, Fineberg DB, Kornfeld SD, Harel NY. Exoskeletal-Assisted Walking for Persons with Motor-Complete Paraplegia. In: Force Sustainment: Rehabilitation, Regeneration and Prosthetics for Re-Integration to Duty; 2013. p. 6–1 – 6–14.

39. Hartigan C, Kandilakis C, Dalley SA, Clausen M, Wilson E, Morrison S, et al. Mobility Outcomes Following Five Training Sessions with a Powered Exoskeleton. Topics in spinal cord injury rehabilitation. 2015;21 2:93–9. doi:10.1310/sci2102-93.

40. Benson I, Hart K, Tussler D, van Middendorp JJ. Lower-limb exoskeletons for individuals with chronic spinal cord injury: findings from a feasibility study. Clinical Rehabilitation. 2016;30:73–84. doi:10.1177/0269215515575166.

41. Kendall MG. A Million Random Digits with 100,000 Normal Deviates. Economica. 1955;22:365. doi:10.2307/2551199.

42. Kirshblum SC, Burns SP, Biering-Sørensen F, Donovan WH, Graves DE, Jha A, et al. International standards for neurological classification of spinal cord injury (Revised 2011). The Journal of Spinal Cord Medicine. 2011;34:535 – 546. doi:10.1179/204577211X13207446293695.

43. Demers L, Weiss-lambrou R, Ska B. The Quebec User Evaluation of Satisfaction with Assistive Technology (QUEST 2.0): An overview and recent progress. Technology and Disability. 2002;14:101–105. doi:10.3233/TAD-2002-14304.

44. Jutai JW, Day H. Psychosocial Impact of Assistive Devices Scale (PIADS). Technology and Disability. 2002;14:107–111. doi:10.3233/TAD-2002-14305.

45. Middleton JW, Yeo JD, Blanch L, Vare VA, Peterson K, Brigden K. Clinical evaluation of a new orthosis, the ‘Walkabout’, for restoration of functional standing and short distance mobility in spinal paralysed individuals. Spinal Cord. 1997;35:574–579. doi:10.1038/sj.sc.3100459.

46. Middleton JW, Sinclair PJ, Smith RM, Davis GM. Postural control during stance in paraplegia: Effects of medially linked versus unlinked knee-ankle-foot orthoses. Archives of Physical Medicine and Rehabilitation. 1999;80(12):1558–1565. doi:https://doi.org/10.1016/S0003-9993(99)90330-1.

47. Prevention and Treatment of Pressure Ulcers/Injuries: Quick Reference Guide 2019 Disclaimer; 2009.

48. Ottenbacher KJ, Hsu YY, Granger CV, Fiedler RC. The reliability of the functional independence measure: a quantitative review. Archives of physical medicine and rehabilitation. 1996;77 12:1226–32. doi:10.1016/S0003-9993(96)90184-7.

49. He Y, Eguren D, Luu TP, Contreras-Vidal JL. Risk management and regulations for lower limb medical exoskeletons: a review. Medical Devices (Auckland, NZ). 2017;10:89–107. doi:10.2147/MDER.S107134.

50. Shimizu Y, Kadone H, Kubota S, Suzuki K, Abe T, Ueno T, et al. Voluntary Ambulation by Upper Limb-Triggered HAL® in Patients with Complete Quadri/Paraplegia Due to Chronic Spinal Cord Injury. Frontiers in Neuroscience. 2017;11. doi:10.3389/fnins.2017.00649.

51. Bowker P. Biomechanical basis of orthotic management; 1993. p. 290.

52. Kerrigan DC, Frates EP, Rogan S, Riley PO. Hip hiking and circumduction: quantitative definitions. American journal of physical medicine & rehabilitation. 2000;79 3:247–52. doi:10.1097/00002060-200005000-00006.

53. Curt A, van Hedel HJA, Klaus D, Dietz V. Recovery from a spinal cord injury: significance of compensation, neural plasticity, and repair. Journal of neurotrauma. 2008;25 6:677–85. doi:10.1089/neu.2007.0468.

54. Guan X, Kuai S, Ji L, Wang R, Ji R. Trunk muscle activity patterns and motion patterns of patients with motor complete spinal cord injury at T8 and T10 walking with different un-powered exoskeletons. The Journal of Spinal Cord Medicine. 2017;40:463 – 470. doi:10.1080/10790268.2017.1319033.

55. Jain NB, Higgins LD, Katz JN, Garshick E. Association of Shoulder Pain With the Use of Mobility Devices in Persons With Chronic Spinal Cord Injury. PM&R. 2010;2. doi:10.1016/j.pmrj.2010.05.004.

56. Ontario Health (Quality). Stance-Control Knee-Ankle-Foot Orthoses for People With Knee Instability: A Health Technology Assessment. Ontario health technology assessment series. 2021;21(11):1–96.

57. Paleg G, Livingstone R. Systematic review and clinical recommendations for dosage of supported home-based standing programs for adults with stroke, spinal cord injury and other neurological conditions. BMC musculoskeletal disorders. 2015;16(358). doi:https://doi.org/10.1186/s12891-015-0813-x.

58. Gorgey AS. Robotic exoskeletons: The current pros and cons. World Journal of Orthopedics. 2018;9:112 – 119. doi:10.5312/wjo.v9.i9.112.

59. Lam T, Noonan VK, Eng JJ. A systematic review of functional ambulation outcome measures in spinal cord injury. Spinal Cord. 2008;46:246–254. doi:10.1038/sj.sc.3102134.

60. Louie DR, Eng JJ, Lam T. Gait speed using powered robotic exoskeletons after spinal cord injury: a systematic review and correlational study. Journal of NeuroEngineering and Rehabilitation. 2015;12(1):1–10. doi:10.1186/s12984-015-0074-9.

61. Shackleton CL, Evans RW, Shamley D, West S, Albertus Y. Effectiveness of over-ground robotic locomotor training in improving walking performance, cardiovascular demands, secondary complications and user-satisfaction in individuals with spinal cord injuries: a systematic review. Journal of rehabilitation medicine. 2019;51(10):723–733. doi:10.2340/16501977-2601.

